# Efficacy of Cerebrolysin Treatment as an Add-On Therapy to Mechanical Thrombectomy in Patients With Acute Ischemic Stroke Due to Large Vessel Occlusion in Anterior Circulation: Results of a three-month follow-up of a Prospective, Open Label, Single-Center Study

**DOI:** 10.1101/2025.02.14.25322320

**Authors:** Jacek Staszewski, Aleksander Dębiec, Stefan Strilciuc, Katarzyna Gniadek-Olejniczak, Renata Piusinska-Macoch, David Balo, George Harston, Adam Stępień, Krzysztof Brzozowski, Piotr Zięcina, Jerzy Narloch, Marek Wierzbicki, Piotr Piasecki

## Abstract

**Background and Aims:** This study hypothesized that Cerebrolysin, a multimodal neuroprotective agent, enhances the efficacy and safety of mechanical thrombectomy (MT) in both acute ischemic stroke (AIS) and recovery stroke phases in selected patients with good collateral status (CTA-CS 2-3) and effective recanalization (mTICI 2b-3).

**Methods:** A single-center, prospective, open-label, single-arm study with blinded outcome assessment of 50 consecutive patients with moderate-to-severe AIS treated with MT ≤6 hours of stroke onset followed by Cerebrolysin (30 ml iv within 8 hours of onset and continued to day 21, first cycle) and in a recovery phase (between 69-90 days, second cycle) compared to 50 historical controls matched by propensity scores. Key outcomes included functional independence (mRS 0–2 at 90 days), safety endpoints, and neurological recovery (NIHSS at 24h and 7 day post MT).

**Results:** Patients receiving Cerebrolysin achieved higher rates of mRS 0–2 at 90 days (68% vs. 44%, p=0.016, OR 2.7, 95%CI 1.2 - 6.1; NNT: 4.2), had reduced risk of secondary ICH (14% vs. 40%, p=0.02; RR 0.37, 95%CI 0.14-0.95), and had lower NIHSS on Day 7 (median [IQR]: 3 [4] vs. 6 [9], p=0.01). There was a significant difference in Barthel Index scores between the Cerebrolysin group and the control group at 30 days (median [IQR]: 77 [32] vs. 63 [50], p=0.03) and at 3 months (86 [22] vs. 75 [29], p=0.01) primarily driven by the increase in the mobility and transfer components. Multivariate analysis identified Cerebrolysin as an independent predictor of favorable outcomes at 3 months (OR 7.5, 95% CI 1.8–30.9), particularly in patients with diabetes (interaction OR 9.6, 95% CI 1.01–92). The overall mortality rates at 30- and 90-days were similar in both groups (2% vs 6% and 8% vs 12%, p>0.1).

**Conclusion:** Cerebrolysin improved functional outcomes at 90 days, accelerated neurological recovery, and reduced complications post-MT in patients with small ischemic core, good collateral circulation and effective recanalization at baseline. These findings warrant further randomized trials to validate its efficacy and explore its long-term benefits.

Registration: URL: https://www.clinicaltrials.gov; unique identifier: NCT04904341

## 1. Introduction

The standards of care for acute ischemic stroke (AIS) due to large vessel occlusion (LVO) have evolved significantly in recent years, driven by advances in evidence-based reperfusion interventions, especially mechanical thrombectomy (MT), and ready access to neurovascular imaging (angio- or perfusion CT [CTA, CTP] or MRI) to guide treatment decisions [^1^]. Despite the improved acute stroke management and postprocedural care and high rates of successful recanalization achieved (over 80%), the rates of functional independence (modified Rankin scale, mRS 0-2) at three months post-procedure still remain suboptimal (30% to 58%) in both early (0-6 hours) and late treatment windows (>6 hours), as reported in clinical trials, registries and everyday practice [^2^,^3^].

Multiple mechanisms have been shown to underlie futile recanalization, i.e. the absence of good outcome after successful endovascular recanalization (TICI 2b-3). These include reperfusion injury due to oxidative stress, activation of inflammatory cascades, calcium overload triggering neuronal apoptosis or necrosis, microvascular failure with “no-reflow phenomenon”, disruption of the blood-brain barrier (BBB) leading to cerebral edema and secondary intracerebral hemorrhage (sICH) [^4^,^5^]. Importantly, a systematic review indicated that the risk of sICH following endovascular treatment (EVT) reaches 40%, and furthermore ICH can also have an impact on functional outcome even if considered asymptomatic by trial definitions [^6^,^7^]. The majority of risk factors for futile recanalization are nonmodifiable [^8^,^9^]. To improve outcome, it will be necessary to explore novel adjunct therapies that target the multiple mechanistic pathways involved in ischemic injury.

Whether adjunctive therapy to MT e.g. neuroprotection could enhance the overall efficacy of EVT in AIS and have significant potential to mitigate MT complications is still not known and it remains an area of current preclinical and clinical research [^10^]. According to the Stroke Treatment Academic Industry Roundtable (STAIR) XI Consortium recommendations, the focus of new studies should be shifted from a neuro- to cerebroprotection or neurovascular protection approach. Different elements of neurovascular unit (NVU) show differential susceptibility to ischemia over differing time scales. The NVU is critical to BBB regulation, cell preservation, and inflammatory immune response during or after AIS [^11^, ^12^]. Therefore, cerebroprotective agents that are multimodal and multicellular, targeting different pathways at time points that are most responsive to treatment might prove more successful [^13^, ^14^].

Cerebrolysin is a biotechnological product which consists of low molecular-weight neuropeptides and free amino acids. It has pharmacodynamic properties similar to those of naturally occurring neurotrophic factors with broad and multifactorial pharmacological properties of cytoprotective potential [^15^,^16^]. It improves cellular survival, inhibits glutamate excitotoxicity, free radical formation, and proinflammatory mediators [^17^]. A recent *in vitro* study showed that Cerebrolysin protects the BBB and has a therapeutic effect on fibrin-impaired cerebral endothelial cell permeability by reducing proinflammatory proteins and by elevating tight junction proteins, therefore reducing hemorrhagic transformation [^18^]. In the CERE-LYSE study, the combination of Cerebrolysin (30 mL/day for 10 days) with r-tPA in AIS was safe, although it did not significantly improve functional outcome in mRS at 3 months. However, in the subgroup with an NIHSS response, significantly more patients had an improvement of 6 or more points in the early recovery phase in the Cerebrolysin group [^19^]. The recent CEREHETIS study demonstrated that early addition of Cerebrolysin to r-tPA significantly decreased the rate of symptomatic ICH (syICH) and early neurological deficit, however it did not influence day 90 functional outcome [^20^]. A small pilot study of Poljakovic et al revealed that Cerebrolysin as an adjunct therapy after reperfusion therapy in patients with futile recanalization decreased mortality rate and risk for sICH [^21^]. Two recent meta-analysis comprising a total of 12 RCT, have recently highlighted the safety and efficacy of combining Cerebrolysin with standardized rehabilitation therapy for motor and neurological function recovery following AIS. These reports led to a positive recommendation for Cerebrolysin use for acute- and post-stroke rehabilitation by the European Academy of Neurology and European Federation of Neurorehabilitation Societies and by other practice national guidelines [^22^,^23^,^24^,^25^,^26^].

We hypothesized that administering Cerebrolysin below 8 hours after stroke onset in carefully selected patients - based on clinical and radiological criteria including moderate to severe stroke, a small baseline ischemic core, good collateral circulation, and significant recanalization following MT - may enhance the effectiveness of MT by triggering cytoprotective effects, mitigating reperfusion injury and the risk of sICH.

## 2. Methods

### 2.1. Study Design and Participants

A prospective, single-center, open-label pilot study with blinded outcome was conducted at the Military Institute of Medicine, Warsaw, Poland, enrolling 50 consecutive AIS patients treated with Cerebrolysin in addition to MT. Historical controls (n=50) matched for age, stroke severity, recanalization grade, bridging r-tPA, occlusion location were used for comparison.

The study protocol was developed in accordance with the Standard Protocol Items: STROBE guidelines for observational studies [^27^]. Detailed study protocol has been described in detail elsewhere [^28^].

Participants for the historical cohort (control group, Ctrl) were retrospectively selected from a large, prospective investigator-maintained registry. The registry has been recruiting since Jan.2018 and is based on a multiprofile and formalized database required for financial settlements with the National Health Fund that includes imaging, demographic, clinical data as well as outcome data (mRS, Barthel Index [BI]) assessed during hospitalization and follow-up at 1-, 3- and 12 months post MT in all patients. The expected proportions of functional independence are 40% and 75% in the historical controls and Cerebrolysin groups, respectively. Such assumptions were based on the results of MT trials that used CTP for patient selection (ESCAPE, SWIFT PRIME, MR CLEAN). Based on the estimated effect size of 35%, a total of 100 patients were required to test the null hypothesis with an a value of 0.05 and a power of 0.8.

Fifty subjects included to the Ctrl group were selected from those treated within a mothership paradigm between June 2018 and May 2021 (n=342, 15%) by the same team of three experienced operators as for CblG and matched one-to-one by the propensity score method (PSM) with those receiving Cerebrolysin [^29^]. The propensity score was estimated using covariable logistic regression model that included ten baseline variables: age, sex, side of stroke and occlusion location (MCA-M1, or MCA-M2), CTA score (2 or 3), pre stroke mRS (0-1), the use of bridging r-tPA and post MT variables: mTICI score (2b, 2c or 3), onset-to-reperfusion time, NIHSS at admission to the Stroke Unit following MT (Fig 1). Propensity score matching was conducted using the nearest neighbor greedy matching algorithm with a maximum caliper width of 0.2. The balance of covariates was evaluated by analyzing standardized mean differences between treatment groups and examining distribution plots for each variable. Patients without suitable matches were excluded to minimize confounders impacting the results.

**Figure 1.**
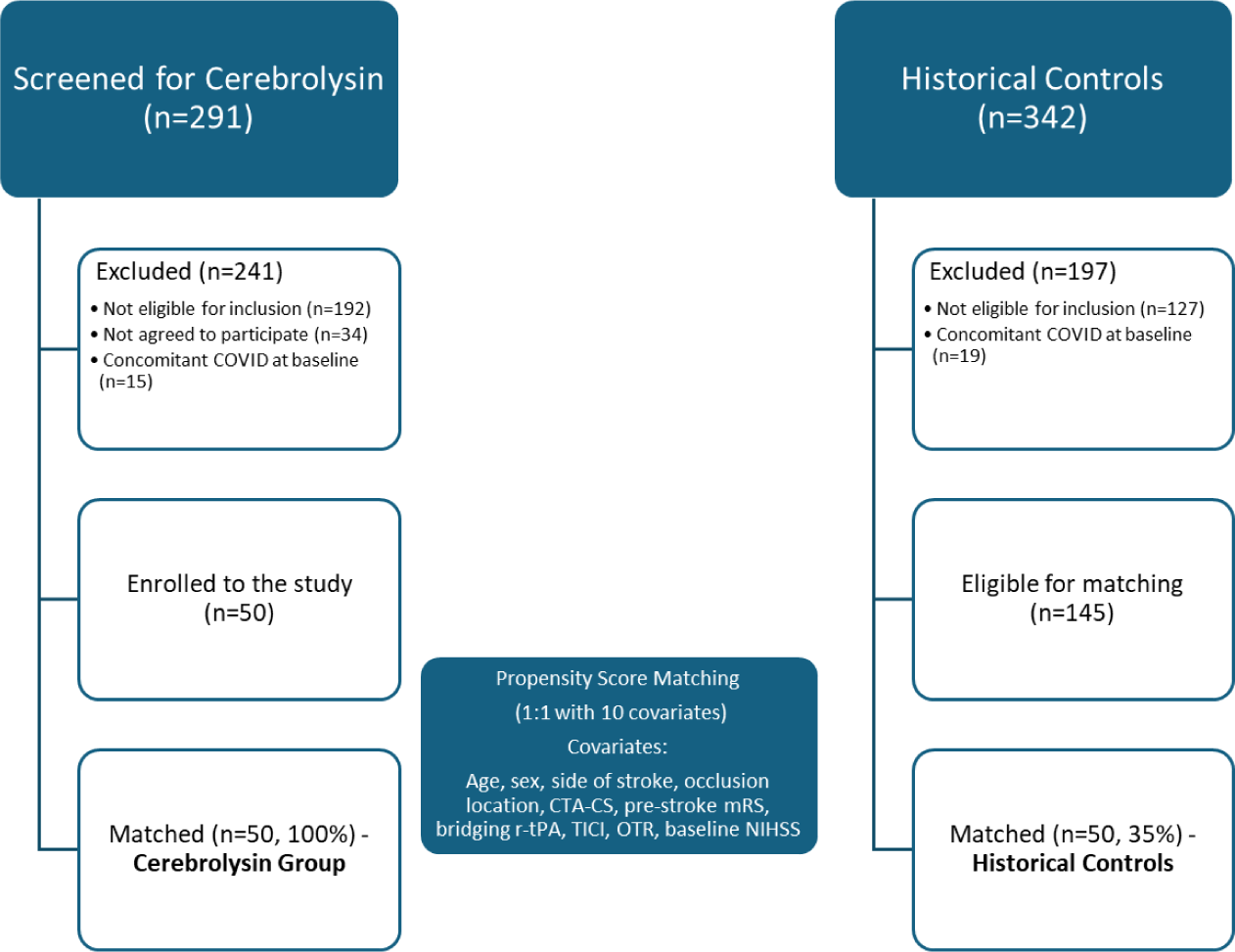
Study flow and analysis of population size after applying exclusion criteria and propensity score matching. OTR – onset to recanalization; TICI – Thrombolysis in Cerebral Infarction grading system; mRS – Modified Rankin Scale; CTA-CS – CT Angiography Collateral Score grading system

The data regarding comparison of CblG and patients eligible for matching is included to the Supplementary Appendix (Figure 1S, Table 1S).

### 2.2 Intervention

All patients received care in the neurointensive care unit and stroke unit care and received stroke treatment according to guidelines. Stroke-trained neurologists administered iv r-tPA to eligible patients prior to MT and patients were jointly qualified for the EVT procedure by a neurologist and an interventional radiologist, according to the AHA/ASA and ESO-ESMINT guidelines [^30^,^1^]. Thrombectomy was performed with any CE-approved thrombectomy device (stent retriever or aspiration thrombectomy or its combination to achieve safe recanalization) at the discretion of the neurointerventionalist [^31^]. All MT procedures were performed by highly skilled neuroradiologists, achieving high rates of successful reperfusion (e.g., mTICI ≥2b in ≥85% of cases) with low complication rates.

Cerebrolysin (30 ml, mixed with 250 ml of Saline/day) was administered intravenously as soon as possible after EVT and within 8 hours of stroke onset, continued for 21 days (during hospitalization or on an ambulatory basis in discharged patients), and repeated from days 69 to 90.

All patients received rehabilitation (physiotherapy, occupational, and speech therapy) at the Stroke Unit following day 1 until discharge and in a Neurorehabilitation Clinic with a minimum of 45min of physiotherapy 5 days a week and according to local standards (the average rehabilitation duration is 3 months post stroke, with the maximum 4 months).

### 2.3 Neuroimaging protocol

The AIS imaging protocol means that all patients assessed for MT receive a noncontrast CT (NCCT), CTA and CTP. Follow-up NCCT is performed routinely at 24 h (all patients) and later in case of neurologic deterioration. To differentiate between sICH and post MT contrast staining in 24h NCCT, in unclear cases we perform follow-up imaging after 24-48 hours with NCCT or MRI with susceptibility-weighted imaging (SWI). Persistent hyperdensity on NCCT or a blooming artifact due to magnetic susceptibility suggests hemorrhage as opposed to contrast staining.

Symptomatic ICH (syICH) was defined as an intracranial hemorrhage accompanied by clinical deterioration (an increase of NIHSS score ≥4 points from baseline) according the ECASS-3 criteria [^32^]. Automated processing of NCCT, CTA, and CTP is routinely performed in every treated patient admitted directly to the hospital using the latest CE-marked version of e-Stroke software (Brainomix, Oxford, UK) as a decision support tool [^33^]. In the current study we used e-Stroke to facilitate adjudication of patient inclusion criteria for prospective patients and historical controls.

### 2.4 Neurological assessments

Neurological assessments were based on a routine evaluation scheme performed in both Cerebrolysin and control patients by a senior neurologist before and following MT (NIHSS_0_), at 24 h post MT (NIHSS_1_), at day 7 (NIHSS_2_, mRS_7D_) and by mRS, BI at 30- (mRS_30D_) and 90 days (mRS_90D_),.

### 2.5 Outcome measures

The primary outcome measure was the proportion of subjects receiving Cerebrolysin experiencing a favorable functional outcome (mRS 0–2) at 90 days following stroke onset compared to the control group.

The secondary objectives were to determine the efficacy of Cerebrolysin as compared to the control group in reducing mortality rates (by 3 months) and the risk of ICH (safety outcomes), improving neurological outcome assessed using NIHSS (at 1 day and 7 day), mRS at 7D and 30D, proportion of patients with mRS 0-1 at 90D, and improving activities of daily living (assessed using BI) at Day 30 and 90. For Day 7 assessments, NIHSS was assigned the maximum score of 42 for deceased patients to reflect the severity of the outcome and maintain consistency in the dataset.

As all stroke patients receiving EVT in our clinic undergo standard neurological evaluation at the predefined time points during hospitalization and after discharge, all clinical and radiological assessments were conducted under blinding conditions where feasible, with assessors blinded to early EVT results and treatment group allocation. All clinical outcomes were obtained through face-to-face consultations during hospitalization or through telephone follow-up after discharge.

### 2.6 Statistical Analysis

The analysis populations included the safety, intention-to-treat (ITT), and per-protocol (PP) groups. The safety population included all patients who received at least one dose of Cerebrolysin compared to all patients in the control group. The ITT population included all patients who met the inclusion criteria, received at least one dose of Cerebrolysin, and had at least observation of the primary efficacy criterion (100% of CblG), regardless of protocol deviations. The PP population included patients from the ITT population who adhered strictly to the protocol without significant deviations (n=41, 82% of CblG used). The primary reasons for protocol deviations included patient refusal to complete the full 21-day Cerebrolysin administration during the first (n = 3) or second treatment cycle (n = 6). Frequently cited reasons for this refusal included a complete sense of recovery and reluctance to extend hospitalization during the ongoing COVD-19 pandemic. Patients with delayed 90D visits during the 2020 lockdown (mean 9 days, SD 4–16 days; n=5) were included in the PP analysis, provided there were no other protocol violations. There was no loss to follow-up or missing data for the clinical secondary outcome measures, except for those related to deceased patients. To assess the robustness of the study results and whether specific baseline characteristics or treatment-related variables are associated with favorable outcomes (mRS 90D 0–2) versus unfavorable outcomes (mRS 90D 3–6), we performed sensitivity analyses including subgroup analyses within the Cerebrolysin group (bridging tPA, ASPECTS <10, AF, anticoagulation, diabetes) and comparisons between the ITT and PP populations. Descriptive statistics and graphs were generated for the ITT population. Statistic analyses were performed by PQStat Software (2024 PQStat v.1.8.6.122. Poznan, Poland).

All patient identification data from both study groups were anonymized to ensure confidentiality. The study was granted by the internal scientific grant by the Wojskowy Instytut Medyczny w Warszawie (No.00589) and reviewed and approved by Institution Review Board of the Wojskowy Instytut Medyczny (No.53/WIM/2020). The patients from CblG provided their written informed consent to participate in this study, for the Ctrl group the consent was not required. This academic investigator-initiated study has been planned to investigate health research questions relevant to everyday practice.

## 3. Results

Between June 2021 and December 2023 we screened 291 patients admitted to our Comprehensive Stroke Center within a mothership paradigm with moderate to severe AIS due to anterior LVO who were treated with MT within 6 hrs of stroke onset and finally included 50 patients (17%) who achieved significant reperfusion following MT (mTICI 2b-3), had good collateral circulation (CTA 2-3) and significant neurological deficit post MT (NIHSS≥5) with cortical signs, who fulfilled other stringent clinical and neuroimaging eligibility criteria (Table 1) and could receive Cerebrolysin within 8 h of stroke onset (Cerebrolysin group, CblG). Severe aphasia or neglect were the most frequent reasons for screening failure as individuals being unable to provide informed consent.

**Table 1.** Eligibility criteria used for Cerebrolysin and Control Groups.

| Clinical Inclusion Criteria | Clinical Exclusion Criteria | Neuroimaging Inclusion Criteria | Neuroimaging Exclusion Criteria |
| --- | --- | --- | --- |
| Age 18–80 years | Other than AIS, serious, advanced, or terminal illness, or life expectancy $\leq 6$ months | CT ASPECTS $\geq 6$ prior to MT | Acute symptomatic arterial occlusions in more than one vascular territory*** |
| Signs and symptoms consistent with the diagnosis of an anterior circulation AIS | Pre-existing medical, neurological, or psychiatric disease that would confound the neurological or functional evaluations** | ICA or MCA-M1 or -M2 occlusion (carotid occlusions can be cervical or intracranial; without tandem MCA lesions) by CTA | Evidence of intracranial tumor (except small meningioma), acute intracranial hemorrhage, neoplasm, or arteriovenous malformation |
| Stroke onset to groin $< 6$ h* | Pregnancy or lactation | Target mismatch profile on CTP (ischemic core volume $< 70$ ml, mismatch ratio $> 1.8$ and mismatch volume $> 15$ ml) | Significant mass effect with midline shift |
| mRS $\leq 1$ prior to qualifying stroke (functionally independent for all ADLs) | Known allergy to iodine that precludes an endovascular procedure | Moderate-to-good collateral status on multiphase CTA ($> 50\%$ MCA territory) | Treatment with another investigational drug within the last 30 days that may interfere with this study's medications |
| Moderate to severe stroke: NIHSS score of $\geq 5$ with the presence of any cortical signs (gaze, visual fields, language, or neglect) | Acute or chronic renal failure with calculated creatinine clearance $< 30$ ml/min/1.73 m<sup>2</sup> or inability to undergo a contrast brain perfusion scan with CT | Effective reperfusion mTICI $\geq 2b$ following MT | Patients with nondiagnostic NCCT or CTP maps |
| Initiation of treatment with Cerebrolysin $< 8$ h following stroke onset **** | Inability to tolerate or comply with study procedures | | |
| Patient has signed the Informed Consent form **** | Any condition that would represent a contraindication for Cerebrolysin administration (e.g., allergy) **** |  |  |

\*Stroke onset is defined as the time the patient was last known to be at their neurologic baseline (wake-up strokes are eligible if they meet the above time limits).
\*\*Examples include Alzheimer's disease, vascular dementia, Parkinson's disease, demyelinating disease, encephalopathy of any cause, or a history of significant alcohol or drug abuse. \*\*\*Examples include bilateral MCA occlusions, or an MCA and a basilar artery occlusion. \*\*\*\* Applies only to Cerebrolysin group

### 3.1. Baseline characteristics and the course of EVT treatment

The primary population and patient characteristics before PSM are shown in Figure 1S, Table 1S of the Supplementary Appendix.

Patients from the CblG and Ctrl groups were well-matched on most baseline variables (Table 2). There were no significant differences in demographics, comorbidities, stroke etiology and severity as well as procedural-related outcomes (OTTICI, number of passes). However, patients from CblG had lower ASPECTS score vs Ctrl (median 9 vs 10p), but both groups did not differ in baseline median mismatch volume (CblG vs Ctrl, 90 ml [IQR 129) and 100 ml [68], p=0,5) and core volume (defined as rCBF<30%: 10 ml [27] and 7 ml [15], p=0,2, respectively). The stroke etiology was evenly distributed between the two groups, with nearly half of the patients experiencing cardioembolic stroke, all attributed to AF. No significant difference in baseline characteristics were found between ITT and PP populations (Table 2S, Supplementary Appendix).

**Table 2.**
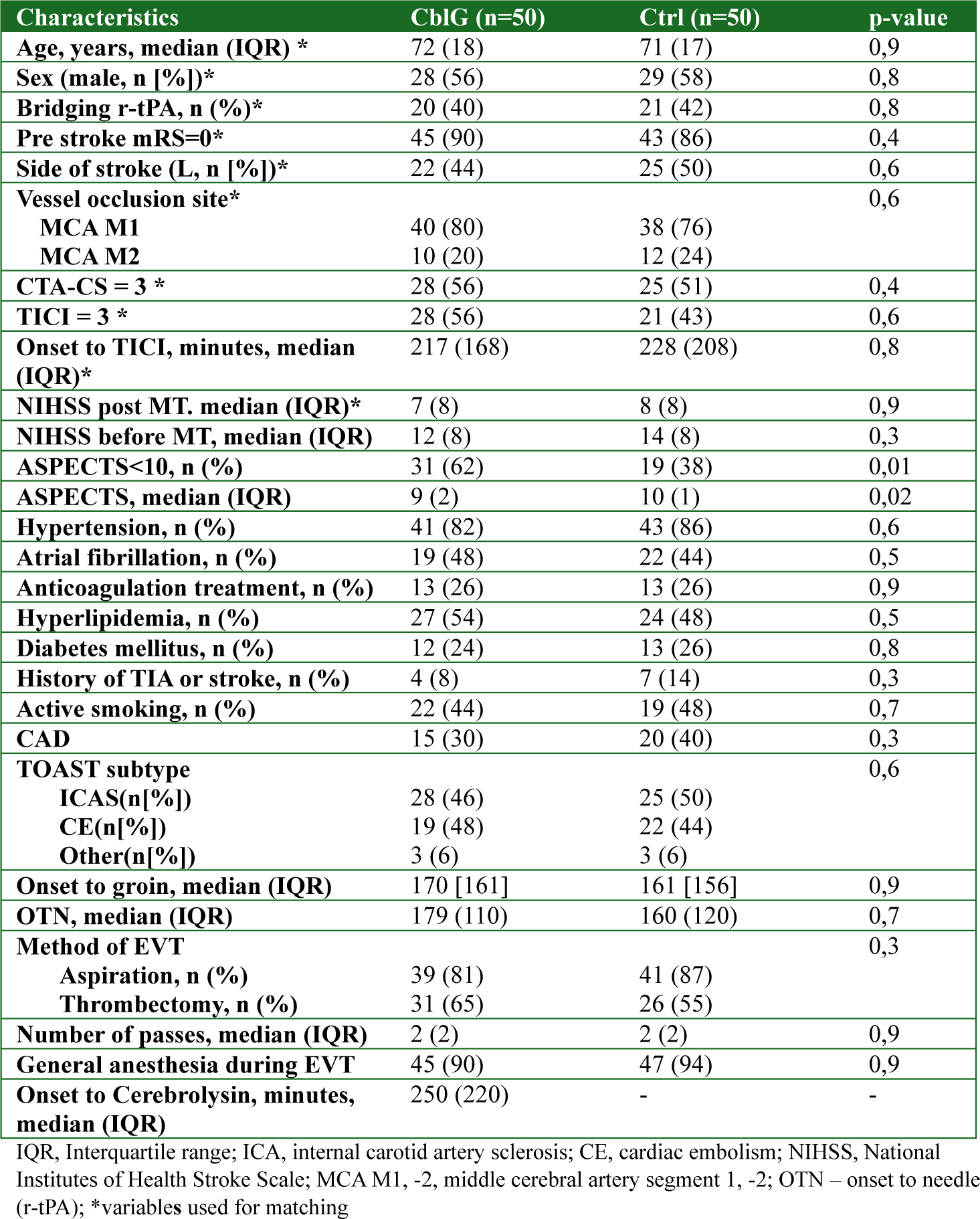
Baseline Characteristics of the Matched Patients.

All EVT procedures were technically successful, and no serious complications occurred during the endovascular procedure (ICH or SAH, dissection, distal embolism, necessity of intracranial stenting).

### 3.2. Outcome

#### 3.2.1 Primary outcome

The study successfully achieved its primary outcome, demonstrating a significant improvement in functional independence at 3 months (measured by mRS 0-2) in the CblG compared to the Ctrl (68% vs. 44%, chi2 p=0.016; OR 2.7, 95%CI 1.2-6.1; NNT: 4.2) (Fig.2). The improvement was higher in a subgroup of patients that received bridging r-tPA (80% vs 47.6%, p=0.03; OR 4.4, 1.1-17.7), remained significant in those with ASPECTS<10 (61.3% vs 26.3%, p=0.02; OR 4.4, 1.3- 15.5) and tended to be significant in patients without any sICH (65.3% vs 34.7%, p=0.07; OR 2.39, 0.9-6.4).

**Figure 2.**
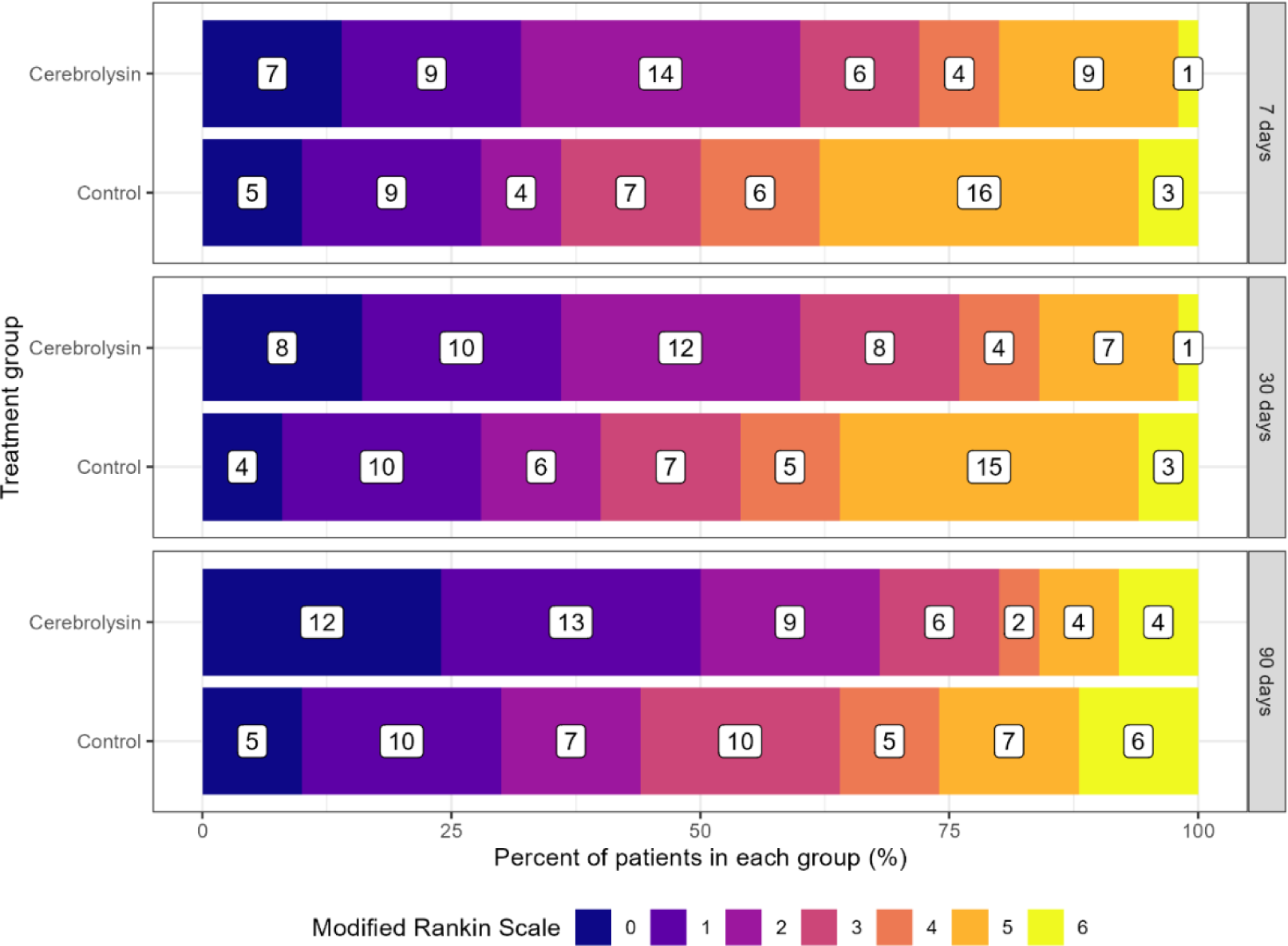
Distribution of 7-, 30-, and 90-day modified Rankin scale scores for patients treated with EVT and Cerebrolysin vs historical controls.

#### 3.2.2. Secondary outcomes

Patients from CblG more often achieved excellent outcome (mRS 0-1) at 3 months compared to controls (50% vs 30%, chi2 p=0.04; OR 2.3,1.02-5.3; NNT 5). Significant differences favoring CblG across the mRS range at 7D (OR 0.79, 0.63-0.99), 30D (OR 0.77, 0.61-0.97) and 3 months (OR 0.78, 0.63-0.97) were also observed (Fig.2). Significantly more patients from CblG compared to Ctrl achieved functional independence at 7- and 30-Days (mRS_7D_ and mRS_30D_ 0-2, respectively 60% vs 36%, p=0.02 and 60% vs 40%, p=0.046). Sixteen patients from CblG (32%) vs 28 (56%) from Ctrl fulfilled the clinical criteria for futile recanalization (mRS 3-6 at 3 months, RR 0.57, 0.276 - 1.184).

There was no significant difference in NIHSS scores at baseline (NIHSS_0_) or at 24 hours (NIHSS_1_) between groups (Fig.3). However, CblG patients performed significantly better than controls on Day 7 post-EVT (NIHSS_2_) (median 3 [4] vs. 6 [9], p=0.01). Patients receiving Cerebrolysin showed quicker and significant improvement between baseline (NIHSS_0_) and Day 1 (NIHSS_1_) (difference in mean ranks: 0.88, p<0.01) and between Day 1 and Day 7 (difference in mean ranks: 0.82, p<0.01). In contrast, the control group did not show statistically significantly improvement (difference in mean ranks: 0.4, p=0.13, and 0.31, p=0.30, respectively).

**Figure 3.**
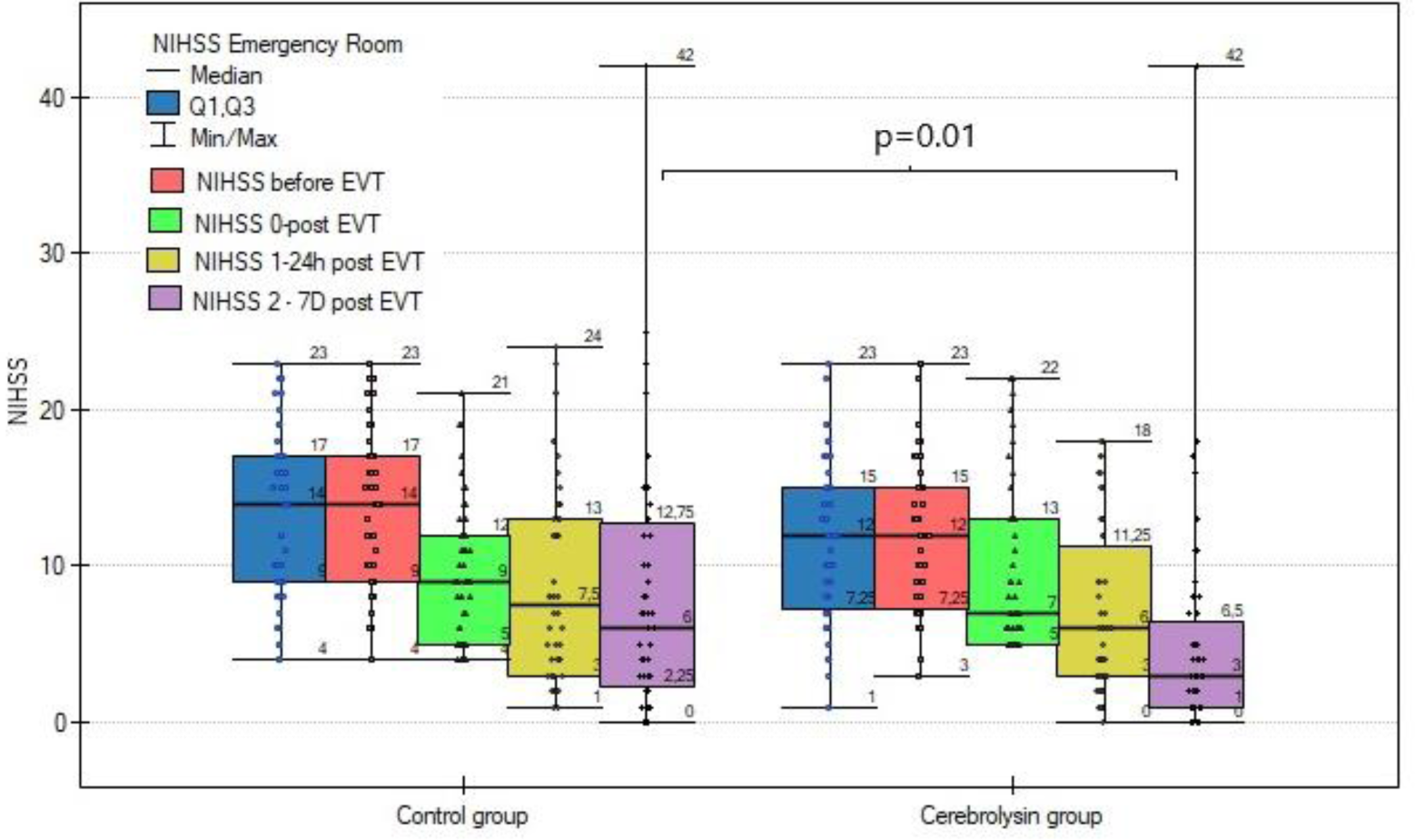
Assessments of NIHSS before EVT, baseline, Day 1 and Day 7 post EVT.

Additionally, there was a difference in median Barthel Index scores between the Cerebrolysin group and the control group at 30 days (77 [32] vs. 63 [50], p=0.03) and at 3 months (86 [22] vs. 75 [29], p=0.01). These improvements were primarily driven by the increase in the mobility and transfer components of BI, which showed the greatest gains in the Cerebrolysin group (+7 points vs. +3 points in the control group, p=0.04).

Significantly more patients from CblG did not need prolonged hospitalization and they were discharged from the hospital directly to home from Stroke Unit or Rehabilitation Department after acute stroke period (44% vs 30%, p=0.02). They also less often required long term institutional care during the 3 months observation period (4% vs 14%, p=0.04).

#### 3.2.3 Safety outcomes

Important differences in safety outcomes were observed between the two groups. Patients in the Cerebrolysin group had a significantly lower risk of any secondary ICH compared to the control group (14% vs. 40%, RR 0.37, 0.14-0.95) and a lower risk of symptomatic ICH (2% vs. 24%, RR 0.08, 0.07-0.097) (Fig. 4).

**Figure 4.**
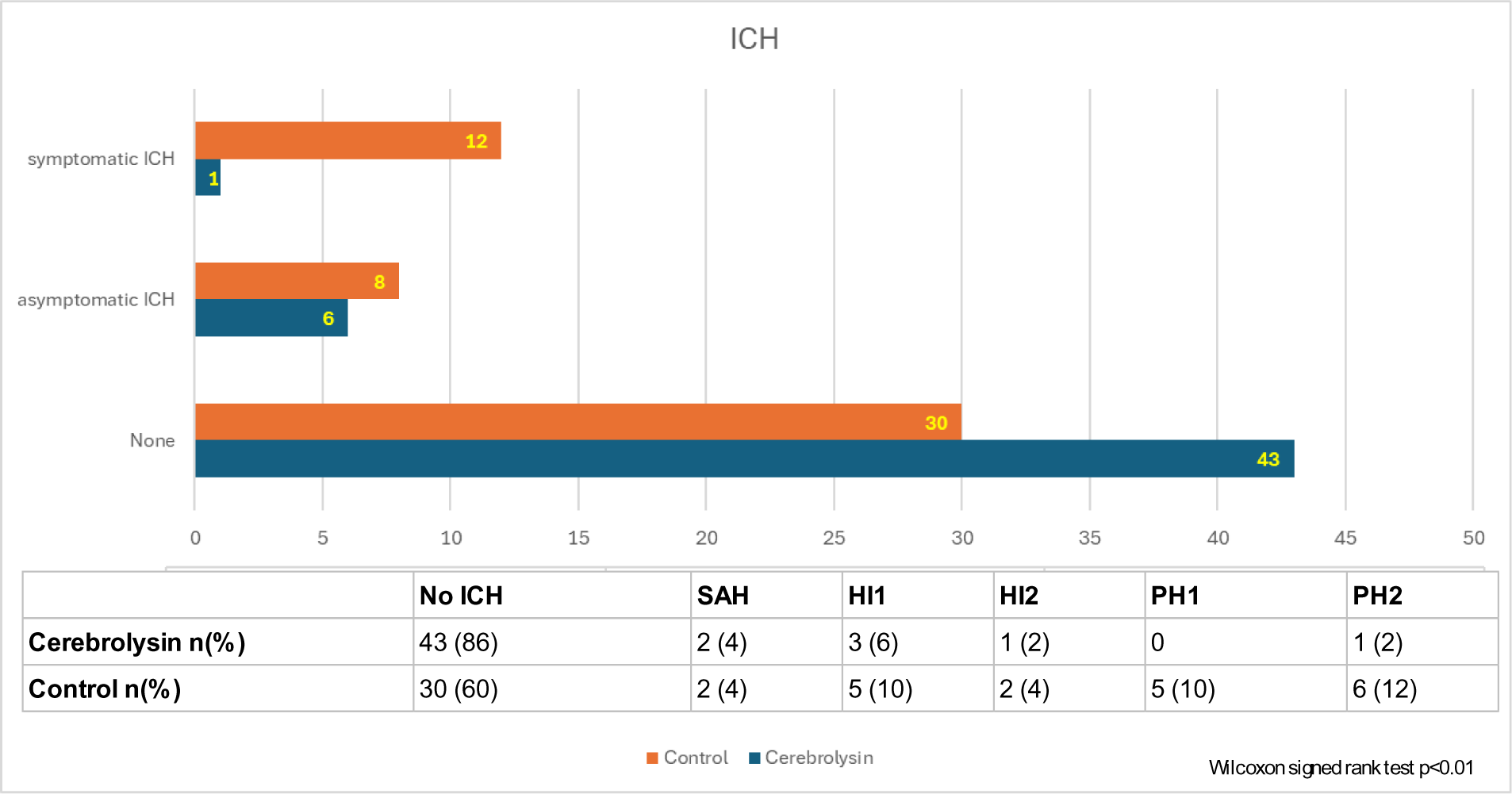
Distribution and type of intracranial hemorrhages according to the study group.

The cumulative 30-day and 3-month mortality rates were similar in CblG and Ctrl groups (2% vs. 6%, p=0.3, and 8% vs. 12%, p=0.5, respectively). In the CblG group, the causes of death were multiorgan and respiratory failure due to COVID-19 (n=2) and stroke-related reasons in two patients (one due to fatal PH2 leading to herniation and one due to recurrent fatal stroke).

Adverse reactions related to Cerebrolysin infusion were reported in 4 patients (8%). These occurred in 3 patients during the first treatment cycle and in 1 patient during the second cycle. All adverse events were mild and consisted of transient superficial venous thrombosis, which developed after median of 10 days of the treatment cycle.

#### 3.2.4. Sensitivity analysis

Analysis within Cerebrolysin group demonstrated key predictors of favorable and unfavorable outcome at 3 month (Table 2S, Supplementary Appendix). Patients with optimal collateral blood flow (CTA-CS 3), complete recanalization (mTICI 3), and diabetes demonstrated significantly higher rates of favorable outcomes (OR 4.6, 1.3–16.5; OR 3.1, 0.9–3.1; and OR 7.1, 0.8–61, respectively). In contrast, patients with cardioembolic strokes (OR 0.2, 0.1–0.9) and those on anticoagulation prior to stroke (OR 0.17, 0.04–0.7) were significantly more likely to experience unfavorable outcomes.

A logistic regression model comparing outcomes between the CblG and Ctrl groups, adjusted for confounders (baseline characteristics and treatment-related variables), identified Cerebrolysin treatment as an independent predictor of favorable outcomes (OR 7.5, 1.8–31) (Fig. 5A). To explore potential modifiers of treatment effects, two models were developed, incorporating clinically relevant variables and their interactions with Cerebrolysin treatment. Model 1 included variables associated with hemorrhagic risk (AF, anticoagulation, bridge r-tPA, ASPECTS<10), while Model 2 focused on metabolic factors (diabetes and concurrent metformin treatment). The analysis revealed that the effect of Cerebrolysin on favorable outcomes was greater in patients with diabetes (OR 9.6, 1.01–92) (Fig. 5B, C). No significant difference was noted when comparing the ITT and PP populations (Table 3S, Appendix).

**Figure 5.**
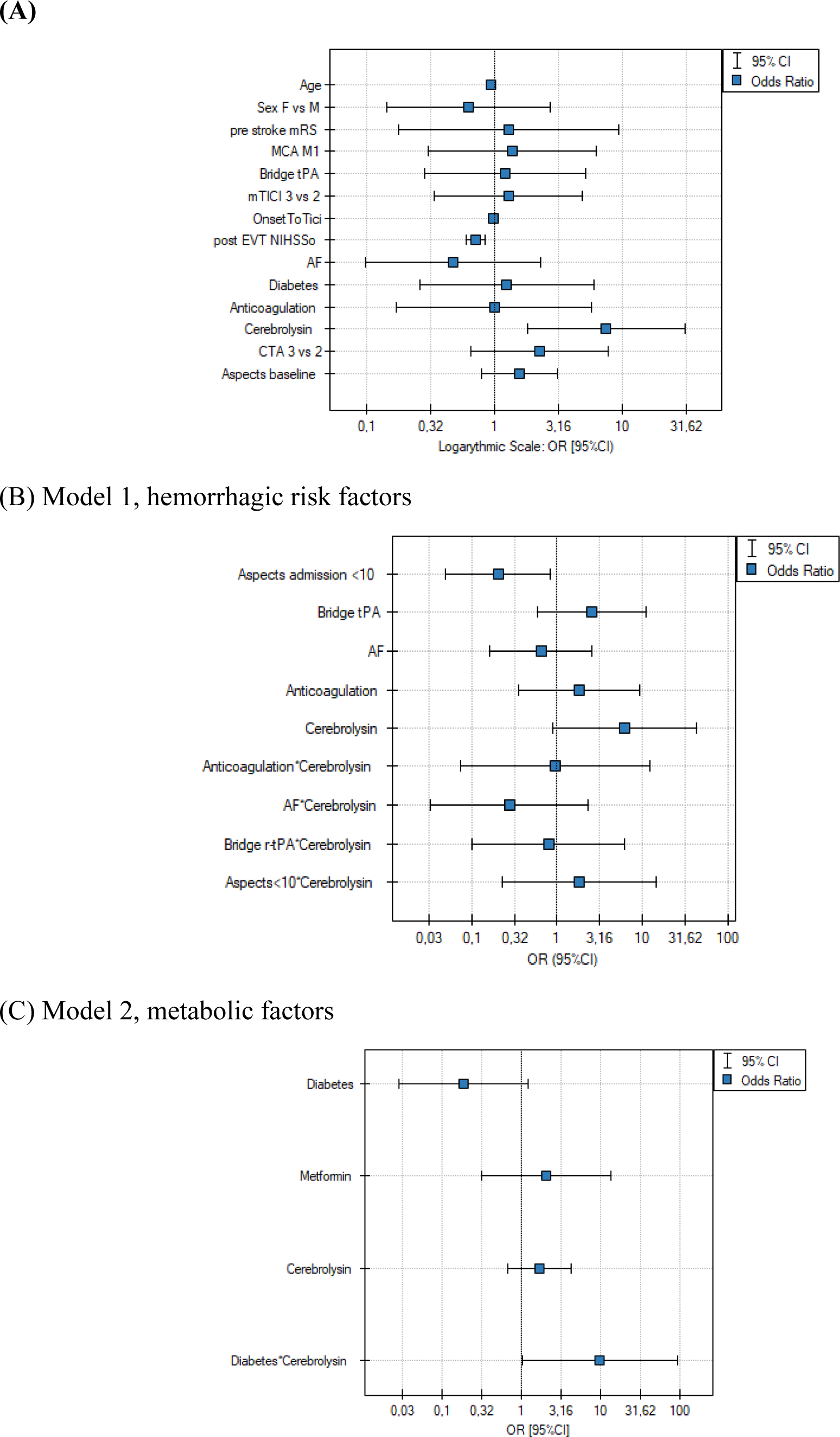
Multivariate logistic regression analysis. (A) Factors associated with achieving mRS 0–2 at 3 months. (B, C) Potential modifiers of the treatment effect of Cerebrolysin.

## 4. Discussion

The findings of this pilot study suggest that Cerebrolysin, administered shortly after successful recanalization following MT in a carefully selected cohort of patients improves both short-term (7 days) and long-term (30-day and 90-day) functional outcomes while reducing the incidence of secondary ICH. To our knowledge, this study is the first to evaluate the effectiveness of combining MT with Cerebrolysin treatment guided by these additional selection criteria.

Functional independence (mRS 0–2, primary outcome) at 90 days was significantly more common in the Cerebrolysin group (OR 2.7, NNT 4.2). The effect was even more pronounced in subgroups treated with bridging r-tPA (OR 4.4) or with baseline ASPECTS <10 (OR 4.4). Additionally, a strong trend toward better outcome was observed in patients without sICH (OR 2.4). Notably, patients receiving add-on therapy with Cerebrolysin demonstrated a lower neurological deficit, as assessed by NIHSS on Day 7. They achieved early functional independence (as evidenced at Day 7, sustained through 30 days post-EVT) and had significantly higher odds (OR 2.3, NNT 5) of excellent outcome (mRS 0–1) at 90 days. There was no significant difference in mortality rates between the groups, however, Cerebrolysin significantly reduced the risk of both symptomatic and asymptomatic ICH by 37%. The treatment demonstrated a favorable safety profile, with no reports of serious adverse events during Cerebrolysin infusion, further reinforcing its safety.

The majority of the treatment effect was consistently observed from Day 7 to Day 90, as demonstrated by mRS outcomes through both ordinal shift and dichotomized analyses. Improvements in neurological status were also evident in NIHSS scores at Day 7 and in BI scores from Day 30 onward. These differences indicate that Cerebrolysin may accelerate and enhance functional independence in patients recovering from neurological events, such as stroke. Although the difference between BI in studied group was not marked, even small increases in BI scores can represent an improvement in independence, such as moving from needing assistance to being independent in specific tasks, reduced caregiver burden and enhanced quality of life.

The early neurological improvement in outcome measures in the Cerebrolysin group compared to controls represents a novel finding. This effect may be attributed to the favorable clinical and radiological profiles of the selected patients, along with beneficial properties of Cerebrolysin on NVU components, such as enhancing the integrity of the BBB, thereby reducing the risk of sICH. Similar findings were reported in the only other published pilot study on the combined use of reperfusion therapy (r-tPA and/or MT) with Cerebrolysin versus controls by Poljakovic et al. [21]. There was a notable trend toward a lower risk of early sICH (13% vs. 38%, p=0.05), a milder neurological deficit (median NIHSS at 7 days: 6 vs. 9, p=0.12), and better functional outcomes at 12 months (mRS 0–2: 70% vs. 48%, p=0.1). Additionally, there was a significant reduction in the 12-month mortality (13% vs. 43%, p=0.03). While we could not confirm a significant impact of Cerebrolysin on 90-day mortality, this may be attributed to patient selection favoring those with a more favorable prognosis (e.g., good collaterals, effective recanalization), the small sample size limiting the statistical power, and the absence of data from a comprehensive 12-month follow-up at the moment.

Significant neurological improvements within the first week after stroke onset, as evidenced by NIHSS and cognitive test scores, were previously documented in the CERE-LYSE trial among patients receiving combination therapy with r-tPA and Cerebrolysin compared to placebo [19]. However, the trial did not show a statistically significant difference in the primary outcome of functional recovery at 90 days. Similarly, the CEREHETIS study demonstrated that early add-on therapy with Cerebrolysin (administered for 14 days) alongside r-tPA was safe and significantly reduced both early neurological deficits (NIHSS on Day 14) and the rate of symptomatic ICH [20]. These effects were likely due to BBB stabilization, attenuation of vasogenic edema, and other cerebroprotective mechanisms, as evidenced by improvements in imaging metrics such as permeability-surface area product, infarct volume, and laterality index. However, no significant impact on functional outcomes at 90 days was observed. The variability in study outcomes, influenced by a more favorable patient profile and outcomes in our study, may also reflect the critical importance of differences in Cerebrolysin administration timing (median 250 minutes in our study vs. 24 hours) and treatment duration (21 days vs. 10–14 days).The early and timely administration of Cerebrolysin in our study (similar to being administered simultaneously with r-tPA in the CEREHETIS study) aligns with the critical therapeutic window for neuroprotection, during which interventions can mitigate excitotoxicity, oxidative stress, and inflammation. In contrast, delayed administration, as seen in other studies (e.g., 1 hour after r-tPA in the CERE-LYSE trial, over 6 hours in trials of Ladurner et al, Heiss et al, and Chang et al) seems to weaken the efficacy of Cerebrolysin on the neurologic functional recovery [^34^, 16, ^35^]. Theoretically, neuroprotective agents should be particularly effective in patients with successful recanalization (resembling a reversible MCA ischemia preclinical model), good collateral flow (ensuring efficient drug delivery to the ischemic core or penumbra), and cortical strokes (which have greater metabolic demand and are more vulnerable to oxidative stress compared to subcortical regions) [^36^]. Cerebrolysin’s neuroprotective effects, including its capacity to mitigate reperfusion injury and stabilize the neurovascular unit by reducing oxidative stress, modulating inflammatory responses, enhancing neural repair mechanisms, and stabilizing the BBB may help reduce these risks as demonstrated in preclinical models [^37^, ^38^]. These considerations were central to the design of the current study and warrant further confirmation in future research.

To facilitate the translation of neuroprotective therapies from research to clinical practice, a novel six-class framework for neuroprotective agents has recently been proposed, tailored to the distinct processes and temporal phases of injury in the reperfusion era including IR injury (Table 4S, Supplementary Appendix [^39^].

An unexpected finding was the favorable interaction between diabetes status and Cerebrolysin suggesting that diabetic patients benefit disproportionately from Cerebrolysin treatment compared to non-diabetic patients and this contradicts typical associations of diabetes with poorer stroke outcomes. This could reflect unmeasured factors, such as better glycemic control or selection bias (the confidence interval is wide, indicating some uncertainty) and warrants further investigation. It is plausible that the observed positive interaction stems from Cerebrolysin enhanced capacity to mitigate the proinflammatory state associated with diabetes, as well as the augmented neuroprotective effects of concurrent metformin therapy [^40^]. Preclinical studies have demonstrated that Cerebrolysin ameliorates cognitive deficits in diabetic models by reducing oxidative stress and modulating inflammatory responses. Additionally, metformin has been shown to possess neuroprotective properties, including the stimulation of endogenous neurorepair mechanisms and the promotion of neural stem cell proliferation and differentiation [^41^].

The safety profile of Cerebrolysin, especially when used in combination therapies, is another critical consideration. Existing data suggest that Cerebrolysin is well-tolerated, with no significant increase in adverse events. Furthermore, a recently published meta-analysis encompassing eight systematic reviews/meta-analyses and six pharmacoeconomic studies confirmed that Cerebrolysin provides cost-effectiveness by improving neurological function recovery, reducing disabilities, enhancing upper limb motor function, and facilitating the recovery of activities of daily living, all while maintaining a safety profile comparable to that of the control group [^42^].

### 4.1. Limitations

While the results are promising, several limitations should be acknowledged. First, the potential impact of selection bias in the study group and the reliance on historical controls cannot be discounted, despite efforts to mitigate this through PSM. Although it adjusts for known confounders, it cannot account for unmeasured variables, which may influence comparability between the study and control groups. Nevertheless, the 44% rate of favorable outcomes at 3 months observed in the historical controls is consistent with the upper range reported in prior EVT trials using CTP for patient selection, such as ESCAPE, SWIFT PRIME, and a subgroup of MR CLEAN, reinforcing the alignment of these findings with established studies. Second, the single-center design may limit the generalizability of the results. Future studies should adopt multi-center, randomized designs to validate these findings. Third, the relatively small sample size may reduce the statistical power to detect additional significant interactions or effects. Fourth, although outcome assessments were blinded, the open-label nature of the study could introduce bias in patient care or evaluations during the intervention phase. Finally, presence and variations in adherence to the second cycle of Cerebrolysin or follow-up protocols could potentially influence outcome measurements. However, no differences in outcomes were noted between the ITT and PP populations, suggesting minimal impact from these variations. Finally, we cannot exclude the impact of COVID-19 on performance in CblG, as the vast majority of enrolments took place during the pandemic.

### 3.2. Strengths

The strengths of the study include the comprehensive outcome assessments with multiple clinically relevant endpoints including functional and neurological status at different timepoints and safety outcomes. This multi-dimensional approach strengthens the study’s relevance to real-world clinical practice. The analysis includes a detailed evaluation of diabetic patients, a population known for poorer stroke outcomes. The use of PSM minimizes selection bias when comparing the CblG with historical controls, enhancing the reliability of the findings. The identification of diabetes as a potential modifier of the treatment effect (interaction OR 9.6) is a significant contribution. Finaly, the study demonstrates a clear benefit of Cerebrolysin in improving functional outcomes and reducing complications, including symptomatic ICH and mortality, providing a strong rationale for further research. These findings provide insights into the practical application of Cerebrolysin in combination with MT, particularly for challenging cases like diabetic patients, where the treatment demonstrated significant benefits. The study’s findings also align with existing preclinical data on neuroprotective effects of Cerebrolysin, strengthening the biological plausibility of its benefits in AIS.

### 3.3 Future directions

Randomized controlled trials are crucial to validate the efficacy of Cerebrolysin as an adjunct to MT. Long-term follow-up studies evaluating functional outcomes and quality of life as well as cognitive functions are also necessary to strengthen the evidence base. Future studies should evaluate the effects of neuroprotective drugs administered as early as possible after stroke onset, including during the prehospital phase or immediately before EVT. These investigations should also consider patients with wake-up strokes and those selected based on tissue-based criteria [^43^]. Further analysis of the collected data in our study is planned, including radiological data (based on a follow-up CT after 1 month) and neuropsychological assessments (battery of aphasia, depression and neglect tests in CblG) as well as outcomes after 12 months for which clearing of the database is currently underway.

## 4. Conclusions

Cerebrolysin used as an adjunct to EVT in AIS patients with good collateral blood flow and successful recanalization, significantly improved both early and long-term (3-month) functional and neurological outcomes and reduced the incidence of secondary ICH. It was especially effective in patients with diabetes highlighting its potential for personalized AIS treatment strategies. However, further randomized, multi-center trials are essential to validate these findings, explore long-term benefits, and define the optimal use of Cerebrolysin in clinical practice.

## Data Availability

Data will be available at a repository after publishing

https://repozytorium.wim.mil.pl/

